# Accelerated 2-Minute Multi-Echo UTE MRI for Simultaneous CT-Like Bone Imaging and Quantitative Short-T2* Mapping

**DOI:** 10.64898/2026.08.18.26360232

**Authors:** Hung P. Do, Mitsuhiro Bekku, Dawn Berkeley, Mark Golden, Shinichi Kitane, Mitsuhiro Uike, Kensuke Shinoda, Ryohei Takayanagi, Hiroshi Takai, Takuma Kawai, Kristen Seballos, Rachel Conley, Kacie Sorfleet, Dustin Devries, Brian Tymkiw, Wissam AlGhuraibawi, Shelton D. Caruthers, Mo Kadbi, Mathew Provencher, Scott Tashman, Charles P. Ho

**Affiliations:** Canon Medical Systems USA 1 Marconi, Suite A/B, Irvine, CA 92618, USA; Canon Inc. 3-30-2 Shimomaruko, Ota-ku, Tokyo 146-8501, Japan; Canon Medical Systems Corporation 1385 Shimoishigami, Otawara-shi, Tochigi 324-8550, Japan; Steadman Philippon Research Institute 181 West Meadow Drive, Vail, CO 81657, USA; The Steadman Clinic 181 West Meadow Drive, Suite 400 Vail, CO 81657, USA

**Keywords:** Multi-echo UTE, UTE-T2* mapping, CT-like bone-weighted contrast, MRI-only MSK imaging

## Abstract

**Purpose:** To determine the feasibility of a 2-minute multi-echo UTE (mecho-UTE) for CT-like bone-weighted contrast and T2* quantification of tissues with short T2/T2*.

**Methods:** Mecho-UTE data acquired from four patients and five healthy subjects were used to assess image quality of the CT-like contrast. All data were reconstructed using conventional gridding (GRID+CONV) and compared with those reconstructed using conjugate gradient SENSE combined with deep learning-based denoising (CG+DLR). Image resolution and sharpness of the CT-like images were assessed using the full width at half maximum (FWHM) and relative edge sharpness (RESH), respectively. Calimetrix UTE-T2* phantom was used to assess the accuracy of T2* quantification of the mecho-UTE sequence.

**Results:** Two-minute mecho-UTE with CG+DLR has similar accuracy (0.37 ± 0.27 vs. 0.67 ± 0.54 ms, p=0.20) and better precision (0.28 ± 0.16 vs. 1.23 ± 0.29 ms, p<0.001) compared to the 5-minute mecho-UTE with GRID+CONV. The 2-minute mecho-UTE with CG+DLR has higher resolution and sharpness compared to the 5-minute scan with GRID+CONV.

**Conclusion:** It is feasible to achieve simultaneous CT-like contrast and T2* quantification of short-T2 tissues in two minutes. When appropriately used, it may simplify logistics, reduce costs, and eliminate radiation exposure risks.

## Introduction

Computed tomography (CT) excels at imaging bone, whereas Magnetic Resonance Imaging (MRI) is superior for imaging soft tissues. For more than three decades, two-dimensional Fast Spin Echo (FSE2D) sequence [1] has been a workhorse of clinical routine musculoskeletal (MSK) MRI because of its efficiency in providing excellent soft-tissue contrast.

Due to the relatively long echo-time (TE), bone and short-T2 tissues (tendons, ligaments, menisci, etc.), although appreciated as signal voids in FSE2D images, cannot be directly visualized or quantified until later disease or damage stages when they appear hyperintense or deformed. Zero echo time (ZTE) and ultrashort echo time (UTE) sequences have been used to provide CT-like bone-weighted contrasts [2–6] but are unable to provide quantitative T2* mapping because of single-echo data. Multi-echo (mecho) UTE, on the other hand, can provide both bone-weighted images and quantitative T2* maps of short-T2 tissues [7–10]. Quantitative T2* map of short-T2 tissues allows them not only to be visualized but also to be quantitatively assessed for tissue health and disease progression. Major drawbacks of UTE and ZTE, i.e., long scan time and low resolution, however, hinder their widespread clinical adoptions despite potential benefits [11–13]. In a review published in 2022, Florkow et al. [11] reported that single-echo ZTE and UTE scan times ranged from 3-10 minutes, in which many have anisotropic resolutions. The shorter the scan time the poorer resolutions reported.

Three-dimensional mecho-UTE sequence utilizes center-out radial k-space trajectory, resulting in a non-Cartesian k-space which cannot be directly reconstructed into images using the inverse Fast Fourier Transform (FFT) algorithm. Alternatively, gridding [14], re-sampling the acquired k-space data into Cartesian grid, is required before applying inverse FFT. Conventional gridding reconstruction with Kaiser-Bessel convolution kernel has been used routinely in clinical settings for UTE and ZTE reconstruction due to its ease of implementation. Gridding is considered a specific type of Non-Uniform FFT (NUFFT) [15]. More advanced implementation of NUFFT [16,17] can be used to reconstruct non-Cartesian k-space data with potential improved accuracy although conventional gridding with appropriate parameters remains the practical method of choice for reconstructing non-Cartesian k-space data [18]. However, when the data is highly under-sampled (e.g., to shorten scan time), both gridding and NUFFT fail to provide diagnostic quality images due to excessive streaking and blurring artifacts [19]. Conjugate gradient sensitivity encoding (CG-SENSE) [20,21], a parallel imaging technique for arbitrary k-space trajectory, can be used to reconstruct under-sampled data with improved image quality.

This work aims to demonstrate the feasibility of a 2-minute three-dimensional 0.8mm^3^ isotropic resolution mecho-UTE using a serial combination of CG-SENSE and Deep Learning-based Denoising Reconstruction (DLR) [22]. The short acquisition time allows it to be added into routine MSK MRI without significant disruption to the clinical workflow. With the advent of Deep Learning-based Reconstruction (DLR), routine MSK MRI can now be acquired in around 5 minutes [23–31] and even as short as 3-minute FSE2D with the knee protocol [32]. Together with DLR-accelerated routine FSE2D MRI, a comprehensive MSK examination can be accomplished in under 10 minutes, where all MSK tissues are directly imaged from soft tissues to bone and those in between (i.e., short-T2 tissues). The MRI-only comprehensive MSK examination would simplify logistics, streamline workflow, reduce costs, and eliminate radiation exposure risks, especially for pediatrics, adolescents, young athletes, pregnant patients, and those who require repeat CT examinations [33–35].

## Materials and Methods

This study was approved by the institutional review board and carried out using 3T MRI scanners. Written consent was obtained from all patients and healthy volunteers.

### Reconstructions

Conventional gridding (GRID) and CG-SENSE are commercially available on the scanner while the prototype deep learning-based denoising reconstruction (DLR) is variant of the commercially available DLR product that was modified for non-Cartesian k-space data. In this study, several sequentially combined reconstructions have been used including GRID+CONV (gridding and then conventional reconstruction filter), GRID+DLR (gridding and then DLR), CG+CONV (CG-SENSE and then conventional reconstruction filter), and CG+DLR (CG-SENSE and then DLR).

### Phantom Experiment

A commercial T2* phantom (Calimetrix LLC, Madison, WI, USA) was used for assessing accuracy and precision of T2* measurements. The phantom contains seven vials with corresponding reference (REF) T2* of 1.1, 1.7, 2.3, 3.3, 4.6, 7.0, and 11.0ms. The phantom was scanned using a three-dimensional mecho-UTE sequence with 5.52-min (R1), 2.76-min (R2), 1.38-min (R4), 0.92-min (R6), and 0.69-min (R8) scan times. The data were reconstructed using GRID+CONV, GRID+DLR, CG+CONV, and CG+DLR. Table 1 lists the sequence parameters used for the phantom experiment.

**Table 1.** Scan parameters for phantom and in vivo experiments using a radial 3D multi-echo UTE. Desired scan times are achieved by reducing or increasing the number of k-space trajectories

| Parameters | Phantom Experiment | *in vivo* Experiment |  |
| --- | --- | --- | --- |
|  |  | Retrospective | Prospective |
| Flip Angle (Degree) | 5 | 5 | 5 |
| TR/TE1/Step (ms) | 20/0.1/2.6 | 12.8/0.096/2.3 | 13.6/0.096/2.5 |
| Number of Echoes | 6 | 4 | 4 |
| Bandwidth (Hz/pixel) | 781 | 781 | 781 |
| FOV (mm<sup>3</sup>) | 190x190x200 | 180x180x200 | 160x160x205 |
| Matrix Size | 256x256x200 | 224x224x250 | 192x192x256 |
| Voxel Size (mm<sup>3</sup>) | 0.7x0.7x1.0 | 0.8x0.8x0.8 | 0.8x0.8x0.8 |
| Scan times (min) | 5.52 (R1); 2.76 (R2); 1.38 (R4); 0.92 (R6); and 0.69 (R8) | 5.8 (R1); 2.9 (R2); 1.9 (R3); 1.4 (R4); and 1.2 (R5) | 5; 3; and 2 |

### Retrospective *in Vivo* Experiment

The 3D mecho-UTE sequence with approximately 5.8-min scan time was scanned on the shoulder of four patients and one healthy subject. The acquired mecho-UTE data was then retrospectively under-sampled with acceleration factors 2, 3, 4, 5 resulting in 2.9-min (R2), 1.9-min (R3), 1.4-min (R4), and 1.2-min (R5) equivalent scan times, respectively. All data was reconstructed using GRID+CONV and CG+DLR for comparison. Table 1 lists the sequence parameters used for the retrospective *in vivo* experiment.

### Prospective *in Vivo* Experiment

The 3D mecho-UTE with scan times of 5-min (n=4), 3-min (n=3), and 2-min (n=3) were performed prospectively on five healthy volunteers. All data was reconstructed with GRID+CONV and CG-DLR for comparison. Table 1 lists the sequence parameters used for the prospective *in vivo* experiment.

Additionally, the mecho-UTE data with 5-min, 3-min, and 2-min scan times were collected on one bilateral hip and one head from two healthy subjects.

### Data Analysis

Non-linear mono-exponential fitting was used to generate the T2* map from the phantom data. Center slice was selected to draw seven circular regions of interest (ROI) on seven vials. Average intensities within each ROI were recorded to assess accuracy of T2* measurement. Standard deviations were used as a surrogate for assessing precision of T2* measurement. Small ROIs were drawn on shoulder tendon and knee ligament from the prospective *in vivo* data to assess T2* measurements across different scan times.

CT-like bone weighted images were generated by using black-and-white reversal of the logarithm of sum of all echo images. Full width at half-maximum (FWHM) of a line profile drawn across a cortical bone was calculated to assess image resolution. Given the same field-of-view (FOV) and matrix size, the image with narrower FWHM means it has higher resolution (less blurring) compared to its counterpart.

The gradient profiles (i.e. successive pixel intensity differences along the drawn line profile) were calculated from the signal intensities of the line profile. Peak gradient amplitudes were then determined from the gradient profiles. The image with higher peak gradient amplitude is considered to have higher image sharpness compared to its counterpart. The relative edge sharpness (RESH) [36] was calculated as a ratio of peak gradient amplitudes of the two images.

A paired Student’s T-test was used to compare mean T2* from different reconstructions and scan times. As recommended by Berchtold et al. [37], linear regression was used to assess reliability, with Bland-Altman and Lin’s concordance-correlation-coefficient for agreement between measured T2* and reference values.

## Results

Fig. 1 shows all possible image contrasts and a quantitative map obtained from routine MSK MRI and an added mecho-UTE sequence. Routine FSE2D (A-E) images provide excellent soft-tissue contrast.

**Fig. 1.**
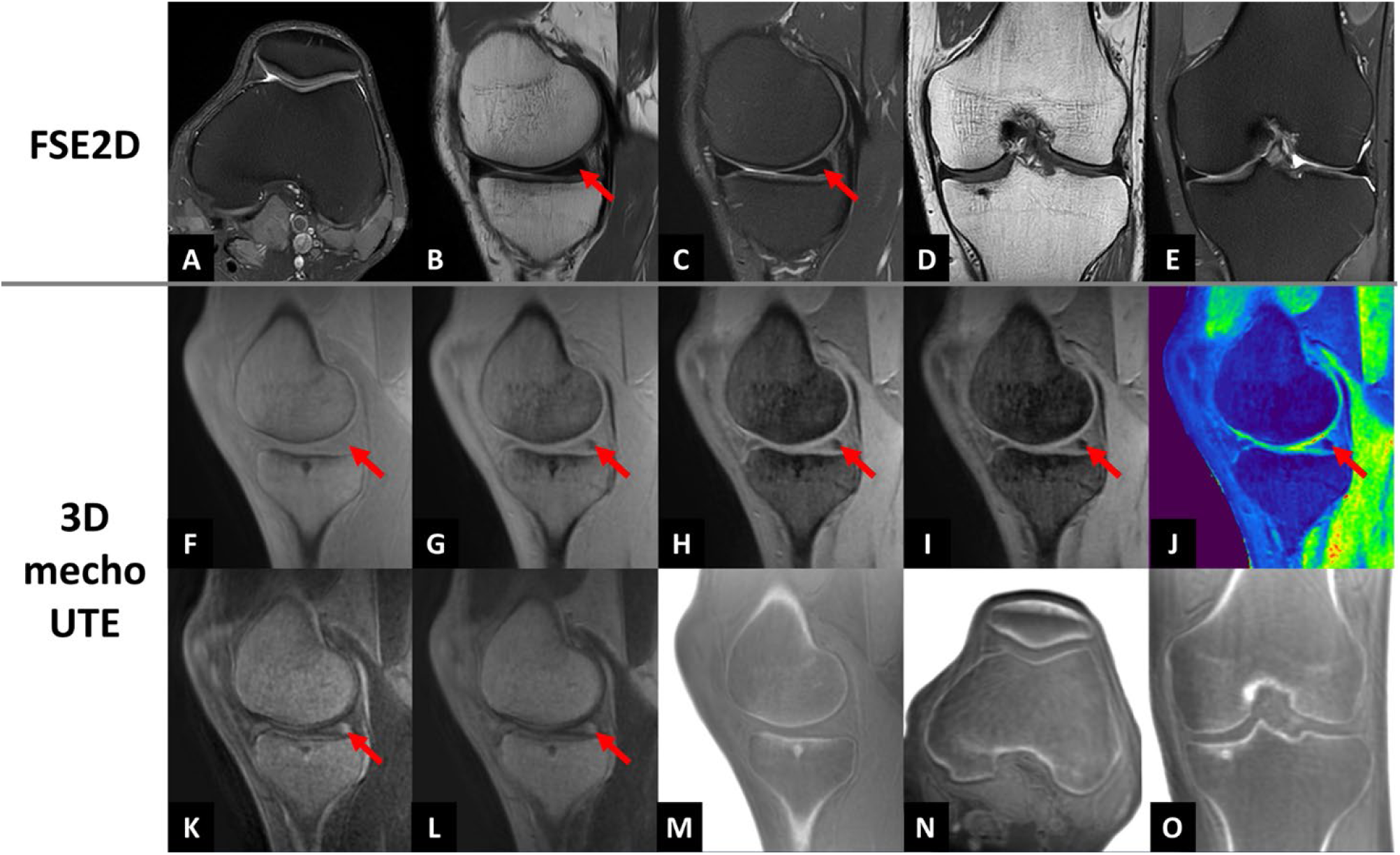
Representative images obtained from the comprehensive MSK protocol. FSE2D images (A-E) provide excellent soft-tissue contrast. The individual echo images (F-I) provide T2*-weighted contrast. T2* map (J) can be generated using non-linear fitting of the multi-echo data to a mono-exponential decay model. To better delineate the short-T2 tissues, echo subtraction images can be generated, as seen in K (TE1-TE2) and L (TE1-TE3). CT-like bone-weighted images can be generated for bone assessment (M: acquired, N and O: reformatted views)

However, due to long TE, cortical bone and short-T2 tissues such as meniscus (arrows), although indirectly appreciated as signal voids, are *invisible* because their MRI signals are significantly decayed by the time of data sampling. Mecho-UTE can provide additional and complementary information to the routine FSE2D images. Individual echo images (F-I) provide T2* weighted images. From mecho-UTE data, T2* map (J) can be generated for quantitative and objective assessment of short-T2 tissues. For visual delineation of short-T2 tissues, echo subtraction images (K: TE1-TE2 and L: TE1-TE3) can be generated. Finally, black-and-white reversal of logarithmic sum of all echo images can be used to generate the CT-like bone-weighted images (M) for bone assessment. The mecho-UTE is 3D isotropic, so it can be reformatted into arbitrary views for assessment of complex MSK anatomies (O, P).

### Phantom Experiment

Accuracy and precision of T2* measurements are shown in Fig. 2. CG-SENSE with and without DLR had better accuracy (A) and precision (B) compared to the gridding reconstruction. More importantly, the 1.38-minute mecho-UTE with CG+DLR (arrows) has similar accuracy (0.37 ± 0.27 vs. 0.67 ± 0.54 ms, p=0.20) and better precision (0.28 ± 0.16 vs. 1.23 ± 0.29 ms, p<0.001) compared to the 5.52-minute mecho-UTE with GRID+CONV.

**Fig. 2.**
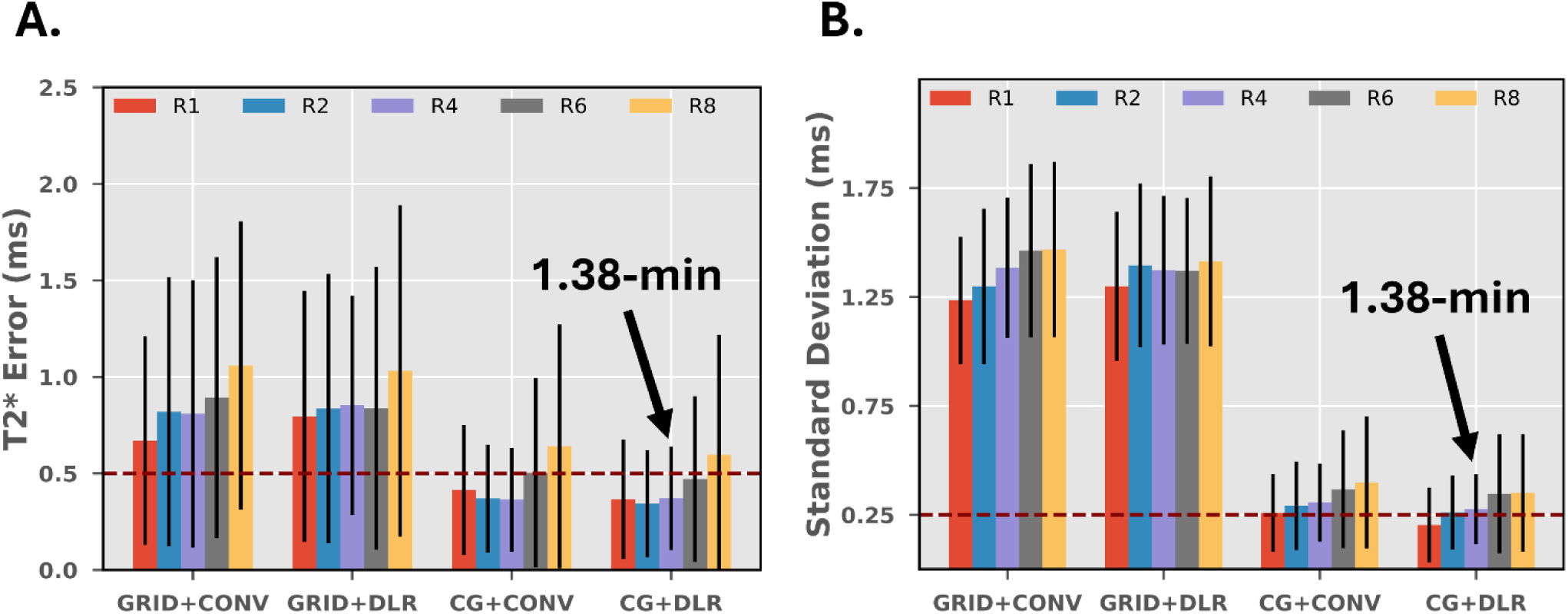
Accuracy (A) and precision (B) of T2* measurements on phantoms. CG-SENSE+DLR (fourth group) provides improved accuracy (A) and precision (B) compared to GRID+CONV (first group). Specifically, the 1.38-minute mecho-UTE (R4) with CG+DLR (arrows) has similar accuracy and better precision compared to the 5.52-minute mecho-UTE (R1) with GRID+CONV. R1, R2, R4, R6, and R8 correspond to 5.52-min, 2.76-min, 1.38-min, 0.92-min, and 0.69-min scan times, respectively

Fig. 3 shows agreement and reliability of the 5.52-min mecho-UTE with GRID+CONV (A, B) and of the 1.38-min mecho-UTE with CG+DLR (C, D) with respect to the reference T2* values. While both scans have sub-millisecond bias, the 1.38-min mecho-UTE with CG+DLR has higher agreement (95% CI: [-1.02, 0.40] ms vs. [-1.92, 1.69] ms) and higher reliability (R-square: 0.992 vs. 0.934) compared to the 5.52-min mecho-UTE with GRID+CONV. Additionally, the 1.38-min mecho-UTE with CG+DLR has higher Lin’s concordance correlation coefficient (0.991 vs. 0.966).

**Fig. 3.**
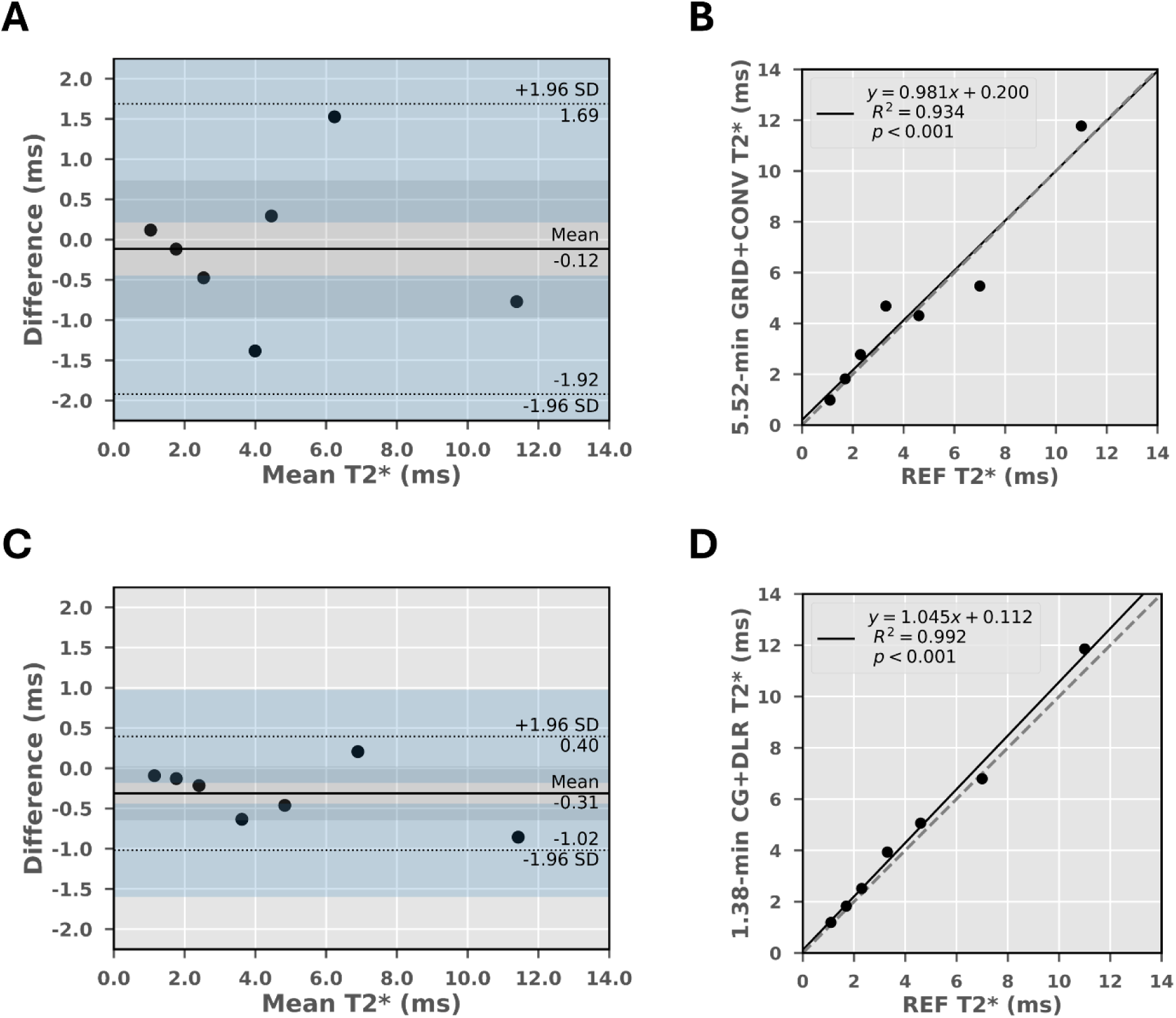
Agreement and reliability of T2* measurements of the 5.52-min mecho-UTE with GRID+CONV reconstruction (A, B) vs. those of the 1.38-minute one with CG-SENSE+DLR reconstruction (C, D). The dashed lines are lines of identity. While both have sub-millisecond bias, the 1.38-min mecho-UTE with CG+DLR (C) has better agreement (95% CI: [-1.02, 0.40] ms vs. [-1.92, 1.69] ms) and reliability (R-square: 0.992 vs. 0.934) compared to the 5.52-min mecho-UTE with GRID+CONV. Additionally, the 1.38-min mecho-UTE with CG+DLR also has a higher Lin’s concordance correlation coefficient (0.991 vs. 0.966)

A picture of the Calimetrix UTE-T2* phantom and a representative T2* map along with the circular regions of interest (ROIs) can be seen in Supplemental Fig. 1. Supplemental Fig. 2 shows T2* maps generated from mecho-UTE data reconstructed with GRID+CONV, GRID+DLR, CG+CONV, and CG+DLR. Supplemental Figs. 3-9 show per-vial T2* and T2* error. Error bars represent the spatial standardization of T2* within a circular region of interest (ROI).

### Retrospective *in Vivo* Experiment

Fig. 4 shows an overview of full width at half-maximum (FWHM) and relative edge sharpness (RESH) assessment with representative images and line profiles. Fig. 4A and 4B show images reconstructed with GRID+CONV and CG+DLR from the same 5.8-min data, respectively, along with line profiles drawn across a cortical bone at the identical location. From the signal intensity of the drawn line profiles shown in Fig. 4A and 4B, FWHM and peak amplitudes of the gradient profiles are determined as shown in Fig. 4C and 4D, respectively. The peak amplitudes are used for RESH calculation. Signal intensity profiles from all retrospectively under-sampled data from the same 5.8-min data reconstructed with GRID+CONV and CG+DLR are shown in Fig. 4E and 4F, respectively. As scan time decreases, the line profiles are broadened (i.e., loss in resolution) with GRID+CONV while they are relatively stable with CG+DLR.

**Fig. 4.**
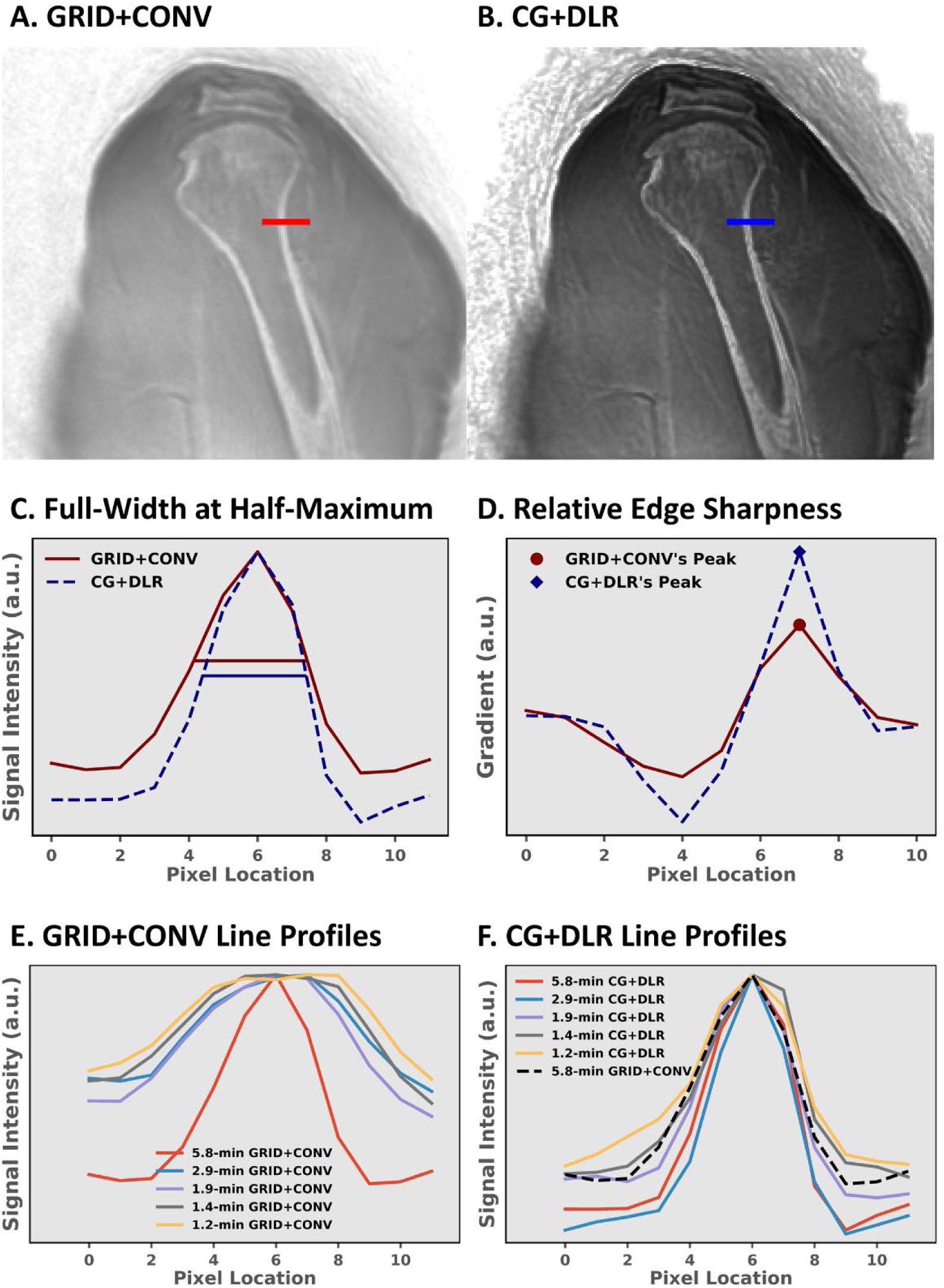
Quantitative metrics for assessing image resolution using full width at half-maximum (FWHM) and image sharpness using relative edge sharpness (RESH). Representative GRID+CONV (A) and CG+DLR (B) reconstructed images with line profiles for calculating FWHM (C) and RESH (D). As scan time decreases, the line profiles are significantly broadened in GRID+CONV reconstructions (E) but relatively maintained in CG+DLR reconstructions (F)

Images from a patient with Bankart bony lesion identified by one of the radiologist-coauthors are shown in Fig. 5. As scan time decreases, GRID+CONV reconstructed images are blurrier (Fig. 5A) while the image quality with CG+DLR reconstruction is relatively stable (Fig. 5B).

**Fig. 5.**
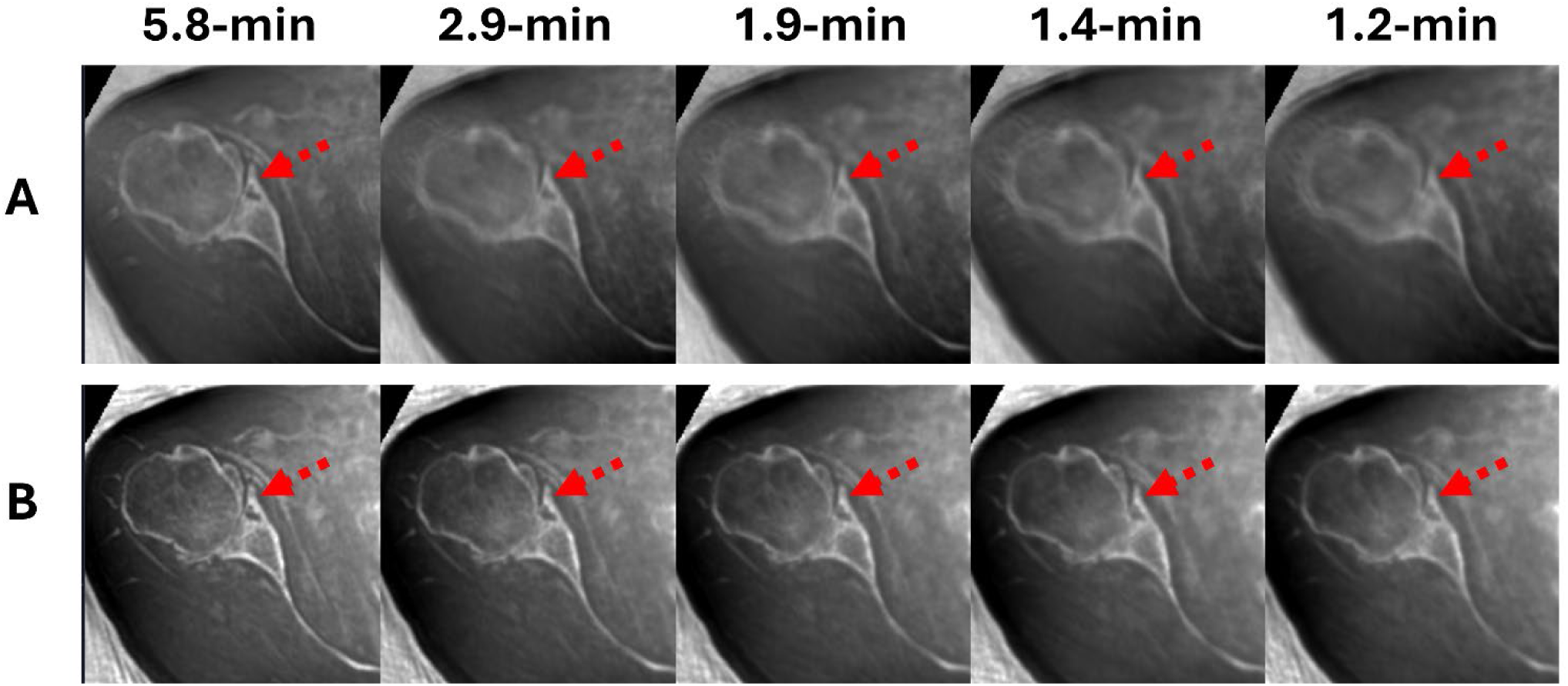
Retrospective study. Bone-weighted images (A, B) generated from a patient with a Bankart bony lesion (red arrows) as identified by one of the radiologist-coauthors. As scan time decreases, the lesion seen in the GRID+CONV reconstructions (A) becomes appreciably blurrier than with CG+DLR (B), in which the sharpness is comparably more stable. All CT-like images are displayed on the same gray scale

To evaluate overall perceived resolution and image sharpness, the FWHM and RESH metrics are summarized for all reconstructed images in Fig. 6. CG+DLR reconstructed images have higher resolution, as indicated by FWHM (Fig. 6A), and image sharpness, by RESH (Fig. 6B), compared to the GRID+CONV counterparts. On average, FWHM from the mecho-UTE with CG+DLR is higher than that from the reference 5.8-min mecho-UTE with GRID+CONV though the difference was not significant (p > 0.24). On the other hand, RESH was significantly higher (p<0.018) for the CG+DLR for the accelerated scan times up to 1.4-min (i.e., 5.8-min, 2.9-min, 1.9-min, and 1.4-min) compared to the reference 5.8-min mecho-UTE scan with GRID+CONV.

**Fig. 6.**
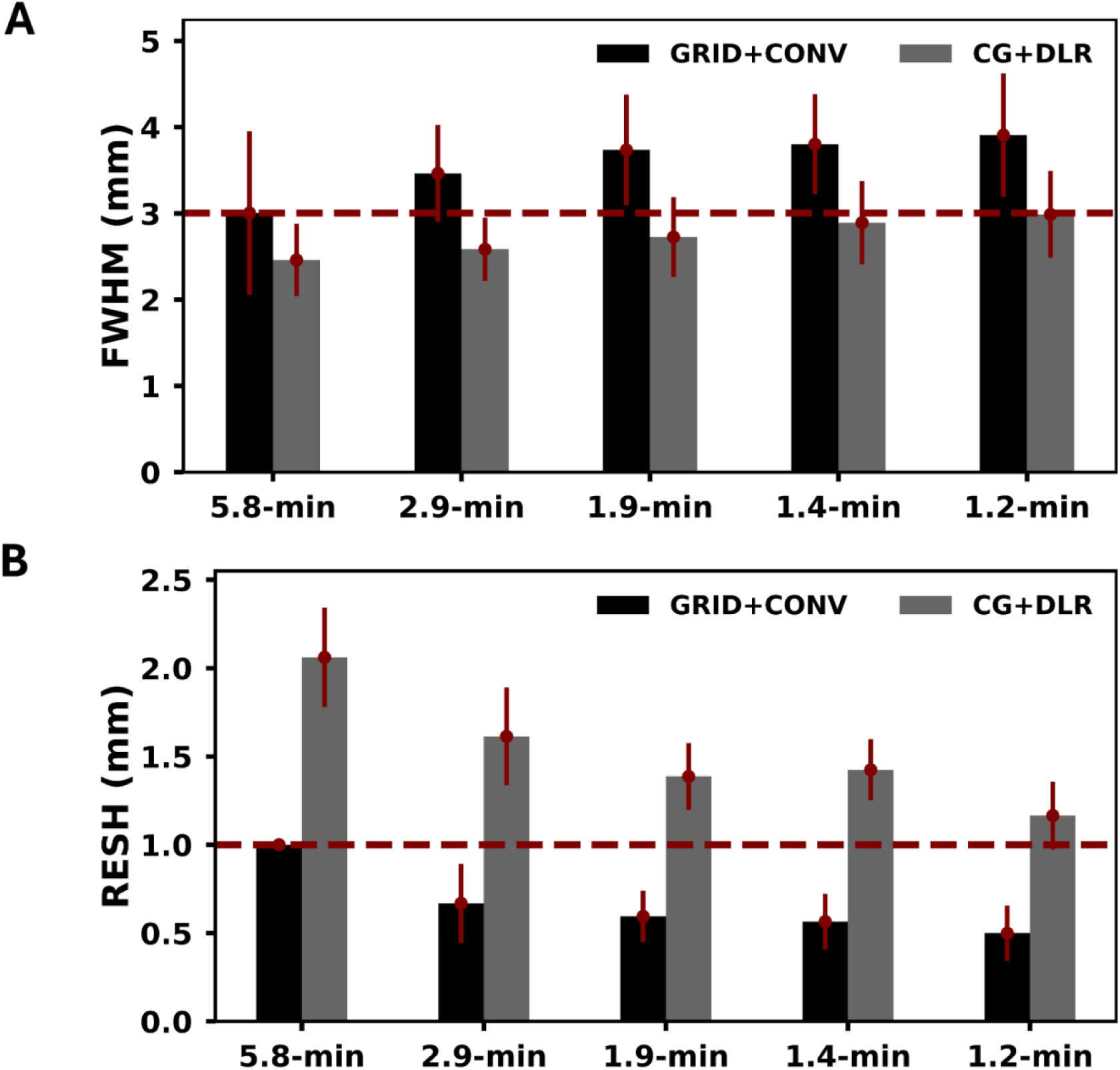
Retrospective study. Compared to GRID+CONV, CG+DLR provides higher resolution (lower FWHM) (A) and higher sharpness (higher RESH) (B). More importantly, up to 1.4-min scan times, CG+DLR has higher resolution and image sharpness compared to the 5.8-min scan with GRID+CONV. Error bars represent standard deviation across subjects

### Prospective *in Vivo* Experiment

For all prospective data, CG+DLR reconstructed images, CG+DLR provides sharper images compared to GRID+CONV. Representative knee images are shown in Fig. 7. CT-like bone weighted images reconstructed from the hip and the brain are reported in Supplemental Fig. 10-11.

**Fig. 7.**
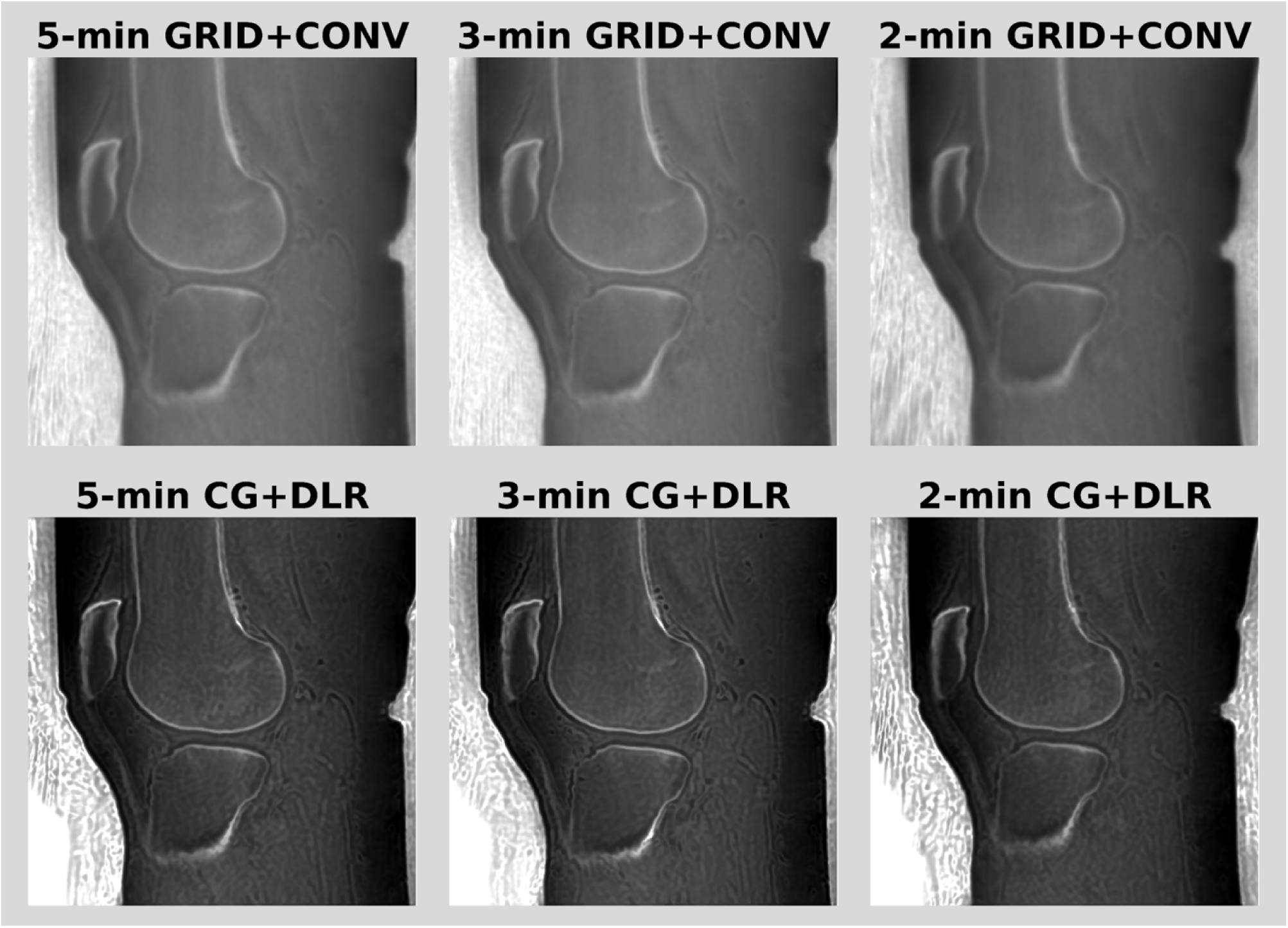
Prospective study. Representative, bone-weighted images from a knee reconstructed with GRID+CONV (top row) and CG+DLR (bottom row). All CT-like images are displayed on the same gray scale

As with the retrospective under-sampling comparisons, the prospectively acquired data demonstrated higher image resolution (Fig. 8A) and sharpness (Fig. 8B) with CG+DLR reconstruction as compared to GRID+CONV. More importantly, 2-min mecho-UTE with CG+DLR has better resolution and sharpness compared to the 5-min mecho-UTE with GRID+CONV.

**Fig. 8.**
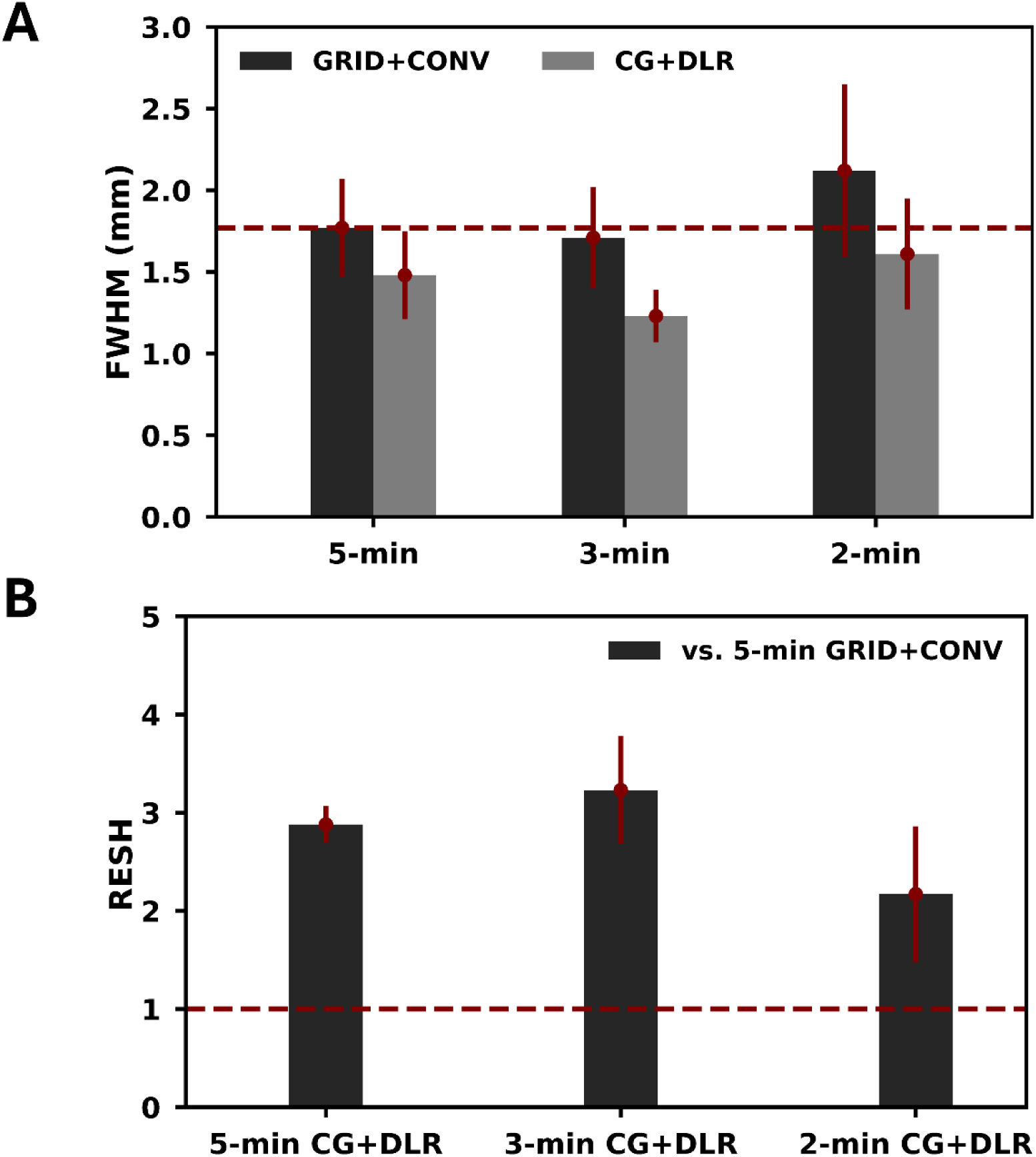
Prospective study. Comparisons of FWHM (A) for resolution and RESH (B) as edge sharpness for the 3 different scan times acquired. Consistently, CG+DLR reconstruction demonstrates better metrics compared to GRID+CONV reconstruction. Notably, the 2-minute mecho-UTE with CG+DLR provides better image resolution and sharpness compared to the 5-minute mecho-UTE with GRID+CONV. Error bars represent standard deviation across subjects

Similar T2* measurements are observed between GRID+CONV and CG+DLR (Fig. 9). CG+DLR shows more consistent T2* values (4.76 ± 1.69, 4.79 ± 1.75, and 4.76 ± 1.74 ms) compared to those from GRID+CONV (5.03 ± 1.47, 5.53 ± 1.19, and 5.72 ± 1.34 ms) across various scan times (5-min, 3-min, and 2-min) likely due to reduced blurring artifacts.

**Fig. 9.**
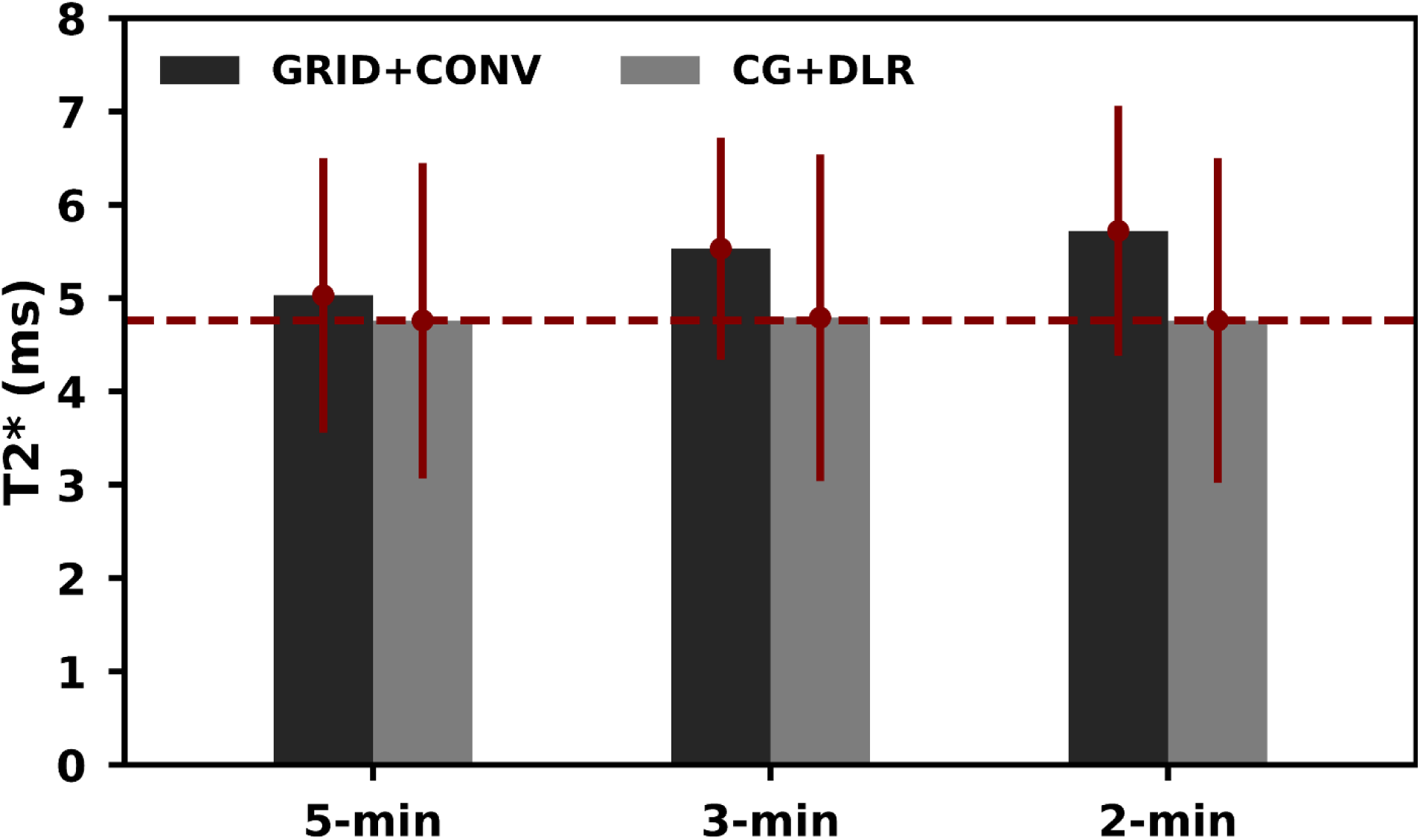
Prospective study. Similar T2* measurements are observed with both GRID+CONV and CG+DLR. A more consistent T2* across scan times seen in CG+DLR is likely due to reduced blurring artifacts

## Discussion

This study demonstrates the feasibility of a 2-minute mecho-UTE reconstructed with CG-SENSE and DLR for simultaneous CT-like bone-weighted contrast and quantitative T2* mapping of short-T2 tissues. Specifically, phantom and human experiments show that 2-min mecho-UTE with CG+DLR provides similar accuracy and better precision for T2* quantification and better resolution and sharpness for CT-like bone-weighted contrast compared to the 5-6-min mecho-UTE with GRID+CONV reconstruction.

Despite potential clinical benefits[12,13], UTE T2* mapping has not been widely used in clinical practice likely due long scan times and lack of standardization. The results reported in literature are heterogenous both in terms of sequence parameters and the measured T2* values. Unlike T2 or T1 mapping where spin-echo MRI can be used as a reference standard for comparison, there is no established reference standard for UTE T2* mapping. Therefore, in this study, we evaluated the T2* accuracy using a commercial Calimetrix phantom consisting of seven vials with T2* ranges from 1.1 to 11ms. Our phantom evaluation showed that the 1.38-min mecho-UTE with CG+DLR has an average T2* error of 0.37 ± 0.27ms [min-max: 0.09-0.86ms]. Short scan time makes mecho-UTE easy to be added into clinical protocol allowing it to be widely evaluated.

Routine MSK MRI can now be acquired in around 5 minutes [23–31] and even as short as a 3-minute knee protocol [32]. When combining with the two-minute mecho-UTE, an MRI-only comprehensive MSK examination can be completed in under 10 minutes. Although it cannot replace CT imaging, when appropriately used, an MRI-only comprehensive MSK examination could simplify logistics, streamline workflow, reduce imaging costs, and eliminate radiation exposure risks.

In broader clinical applications, the 2-minute mecho-UTE can be feasibly added to any abbreviated MSK protocol, wherein focused sequences were selected to provide optimal diagnostic yield per unit of scan time, tailored to answer a specific clinical question, leading to “CT-like” throughput capability with less than 5 minutes total scan time. In a recent study, Huang et al. [38] showed that US Medicare/Medicaid reimbursements in 2025 for orthopedic MRI (upper and lower extremities without contrast) were reduced by 77% compared to their peak in 2004. This study underscores the need for implementing an abbreviated MSK MRI examination that can provide the highest clinical value in the shortest possible time. Additionally, if the MRI-only protocol is cost-effective enough, it may be used as the first-line imaging when appropriate [39], especially to avoid the necessity of subsequent MRI examinations [40].

Short-T2 tissues such as ligaments, menisci, and tendons can be indirectly appreciated from routine MRI as signal voids, which often appear hyperintense or deformed at later stages of damage or disease. One potential benefit of mecho-UTE is that it can provide not only better visualization of short-T2 tissue in the early echo images or in the echo-subtraction images, but also quantitative assessment through the T2* map. Studies have shown that T2* elevation is associated with the severity of disease progression and degeneration [8,10,41,42]. Accurate T2* mapping may allow early and longitudinal characterization of the disease progression and recovery.

ZTE is frequently used for CT-like bone-weighted imaging, however, it has not been reported to simultaneously provide CT-like bone-weighted contrast and T2* map of short-T2 tissues in clinically relevant scan time. Multi-echo Gradient Echo (mecho-GRE) has also been shown to generate CT-like bone-weighted images [43], however, due to longer TE than UTE, it is also unable to accurately generate T2* map for short-T2 tissue.

Though not evaluated in this study, it is worth noting that the multi-echo UTE can be used to further enhance visualization of short-T2 tissues such as tendons, menisci, and ligaments by utilizing optimized weighted subtraction [44] instead of directly subtracting two echo images.

For more than three decades, FSE2D has been an indispensable sequence of MSK MRI. The sub-2-minute mecho-UTE can be conveniently added into any MSK protocol without any major disruption, hence can provide immediate impact. Although mecho-UTE does not replace existing FSE2D or CT but, when appropriate, can be added into any MSK protocol to provide complementary information with added benefits[7,11–13].

This feasibility study has several limitations. First, test-retest repeatability and reproducibility were not fully performed. Second, this study included a small number of healthy subjects and patients and without head-to-head comparison with CT as a gold standard for diagnostic confidence. Furthermore, future studies are warranted to validate the proposed protocol on large-scale multi-site clinical trials where diagnostic interchangeability, time-efficiency, cost-effectiveness, and safety against the standard of care of using both MRI and CT would be evaluated.

## Conclusion

CG-SENSE and DLR enable 2-minute mecho-UTE for simultaneous CT-like bone-weighted imaging with accurate T2* quantification of short-T2-tissues. Together with a 3-to-8-min routine clinical MSK protocol, a comprehensive MSK examination can be performed in under 10 minutes. When appropriately used, the MRI-only protocol would simplify logistics, streamline workflow, reduce costs, and eliminate radiation exposure risks, especially for pediatric, adolescent, pregnant patients, and those that require repeated CT examination.

## Funding

This study was funded by Canon Medical Systems USA.

## Data availability

The data that support the findings of this study are not publicly available because of ethical, legal, and privacy restrictions and therefore cannot be shared.

## Ethics approval

This study was performed with approval from the Institutional Review Boards of the Steadman Clinic and Canon Medical Systems USA.

## Informed consent

Written informed consent was obtained from all participants.

## Conflict of interest

H. P. D., D. B., B. T., W.A., and M. K. are employees of Canon Medical Systems USA. M.B., M.G., S.K., M.U., K.S., R.T., H.T, K.T., and S.C. are employees of Canon Inc., Japan. Additionally, C.P.H. is a consultant for Smith & Nephew and to Miach Orthopedics, not relevant to this study. The other authors declare that they have no conflict of interest.

## Abbreviations

Mecho-UTE: Multi-echo Ultrashort Echo Time
ZTE: Zero Echo Time
DLR: Deep Learning-based Denoising Reconstruction
CG-SENSE: Conjugate Gradient Sensitivity Encoding
MSK: Musculoskeletal
FWHM: Full-Width at Half-Maximum
RESH: Relative Edge Sharpness
FSE2D: Two-dimensional Fast Spin Echo;

## Supplemental Figures

**Supplemental Fig. 1.**
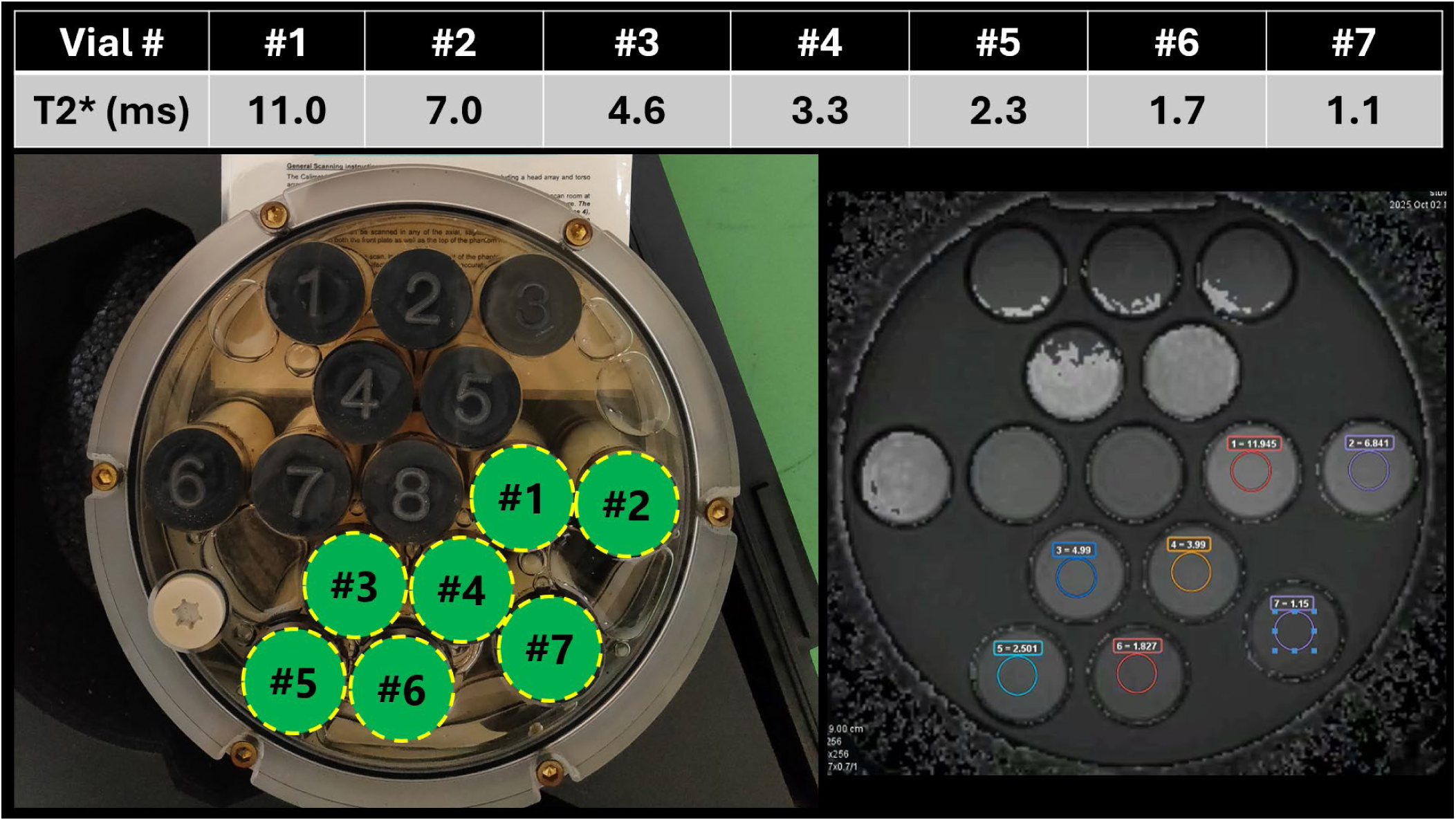
Calimetrix UTE-T2* phantom (left) with T2* values on the table. A representative T2* map (right) with regions of interest (ROIs).

**Supplemental Fig. 2.**
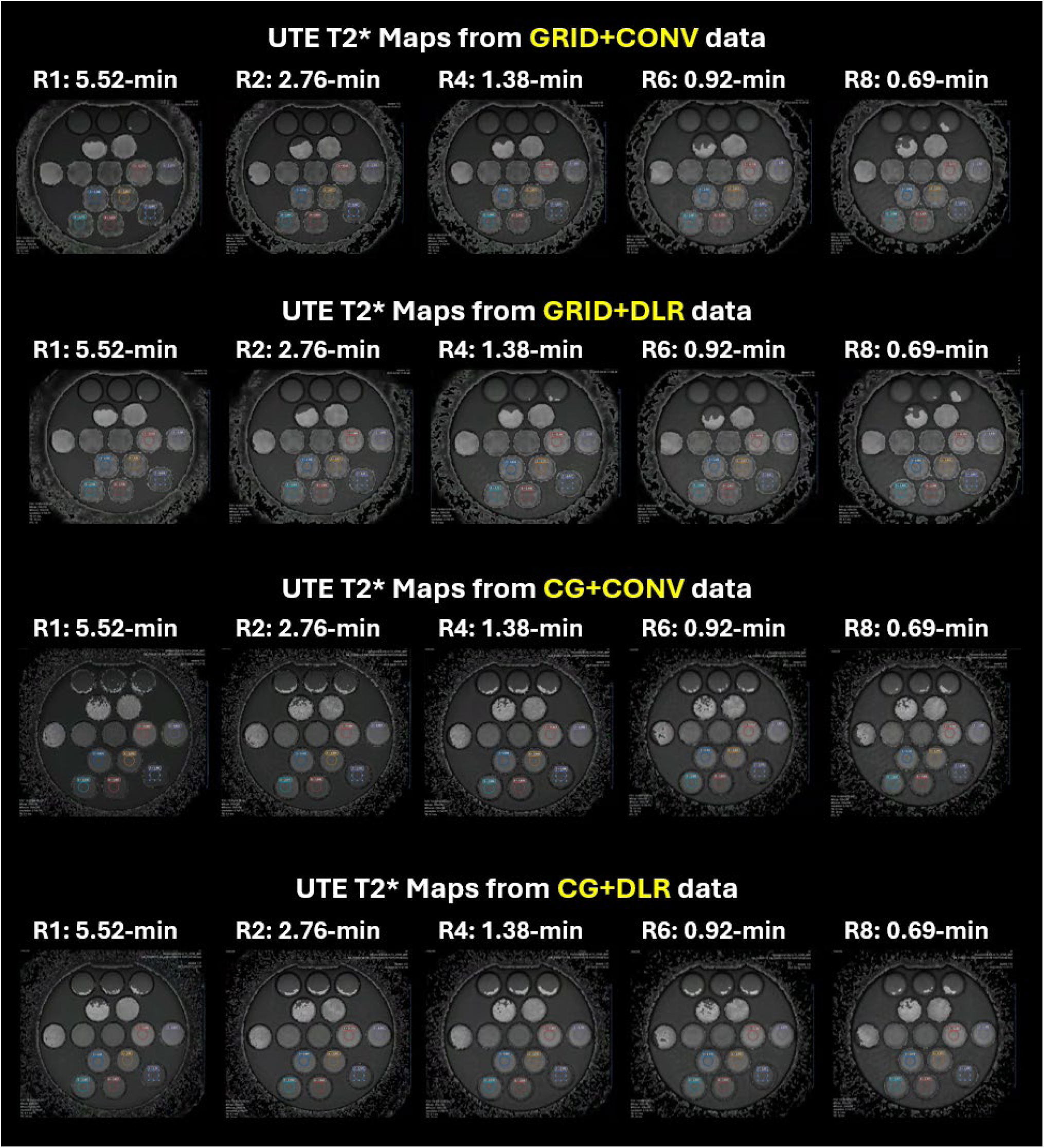
T2* maps generated from mecho-UTE data reconstructed with GRID+CONV (first row), GRID+DLR (second row), CG+CONV (third row), and CG+DLR (fourth row).

**Supplemental Fig. 3.**
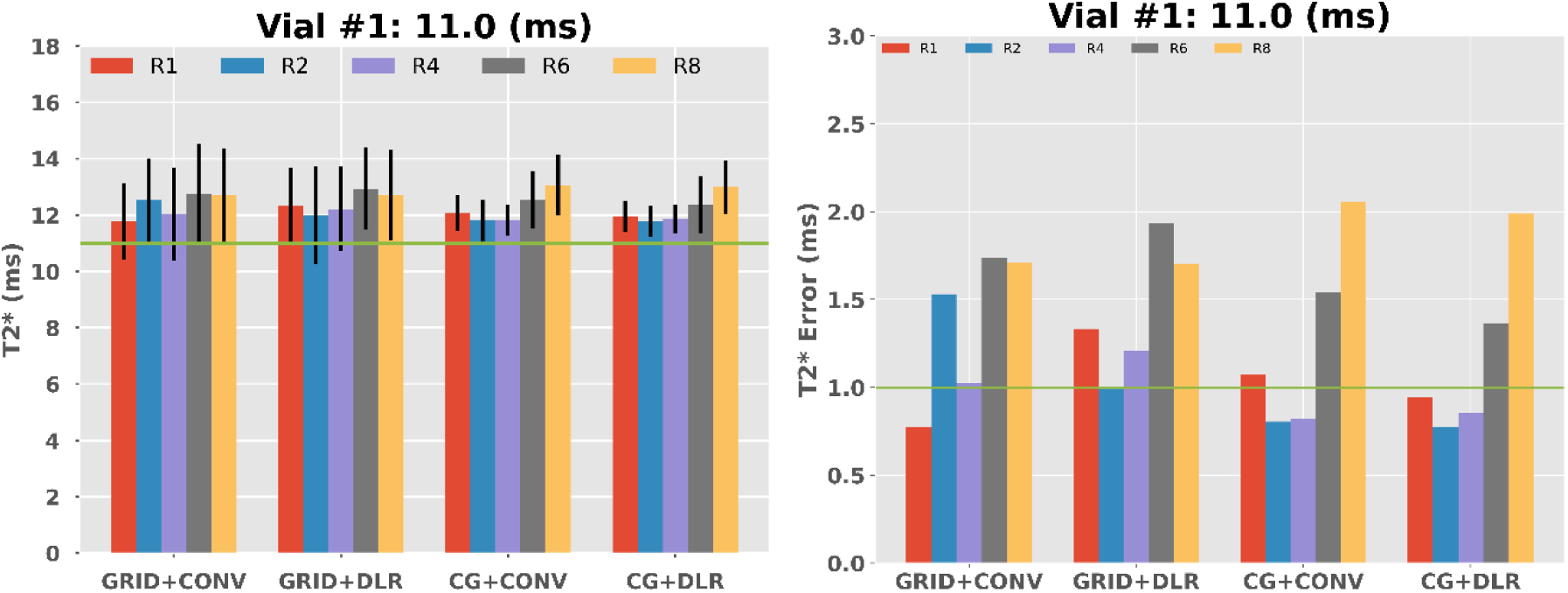
T2* (left) and T2* error (right) measured from **vial #1** with different accelerations and reconstruction methods. Error bars represent the spatial standard deviation of T2* values within an ROI.

**Supplemental Fig. 4.**
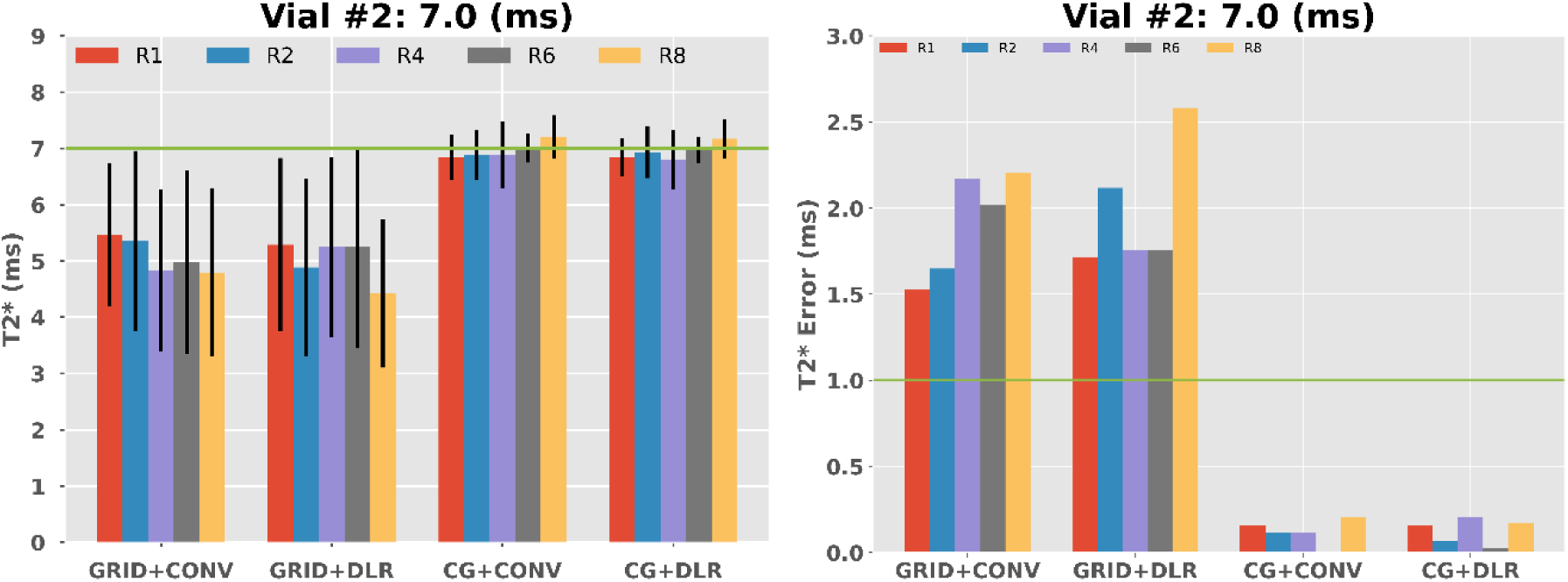
T2* (left) and T2* error (right) measured from **vial #2** with different accelerations and reconstruction methods. Error bars represent the spatial standard deviation of T2* values within an ROI.

**Supplemental Fig. 5.**
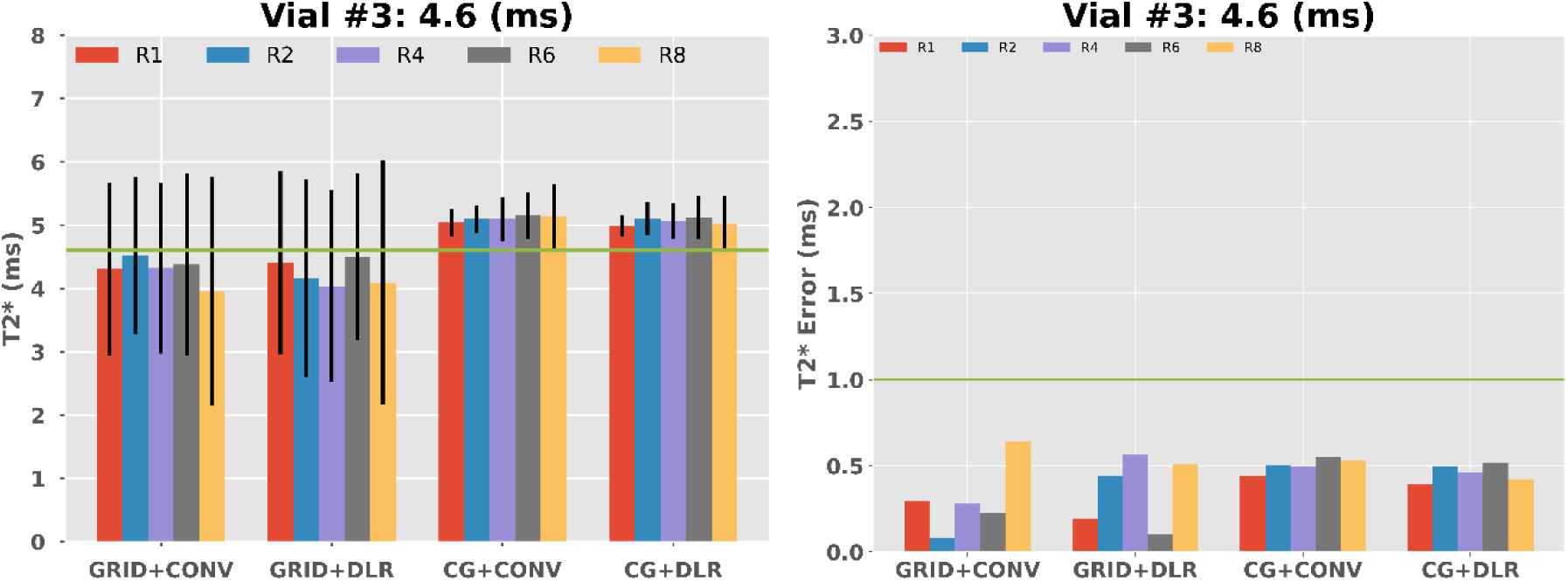
T2* (left) and T2* error (right) measured from **vial #3** with different accelerations and reconstruction methods. Error bars represent the spatial standard deviation of T2* values within an ROI.

**Supplemental Fig. 6.**
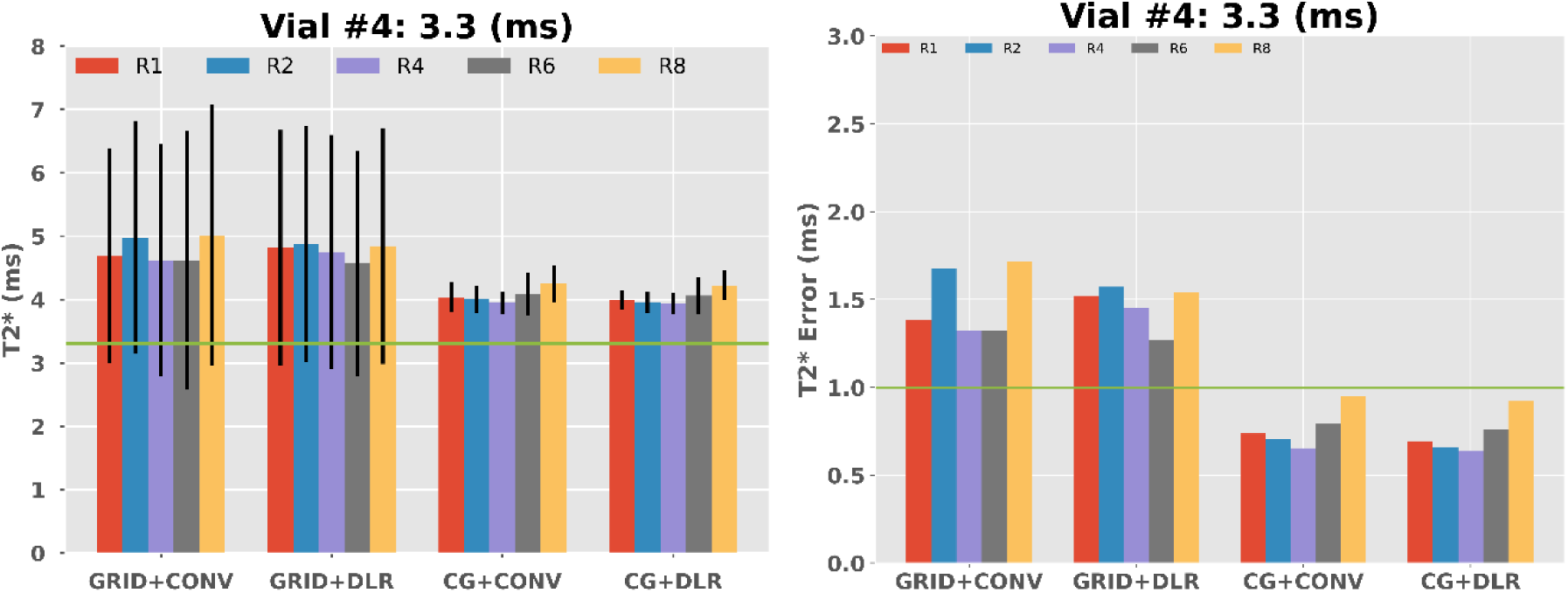
T2* (left) and T2* error (right) measured from **vial #4** with different accelerations and reconstruction methods. Error bars represent the spatial standard deviation of T2* values within an ROI.

**Supplemental Fig. 7.**
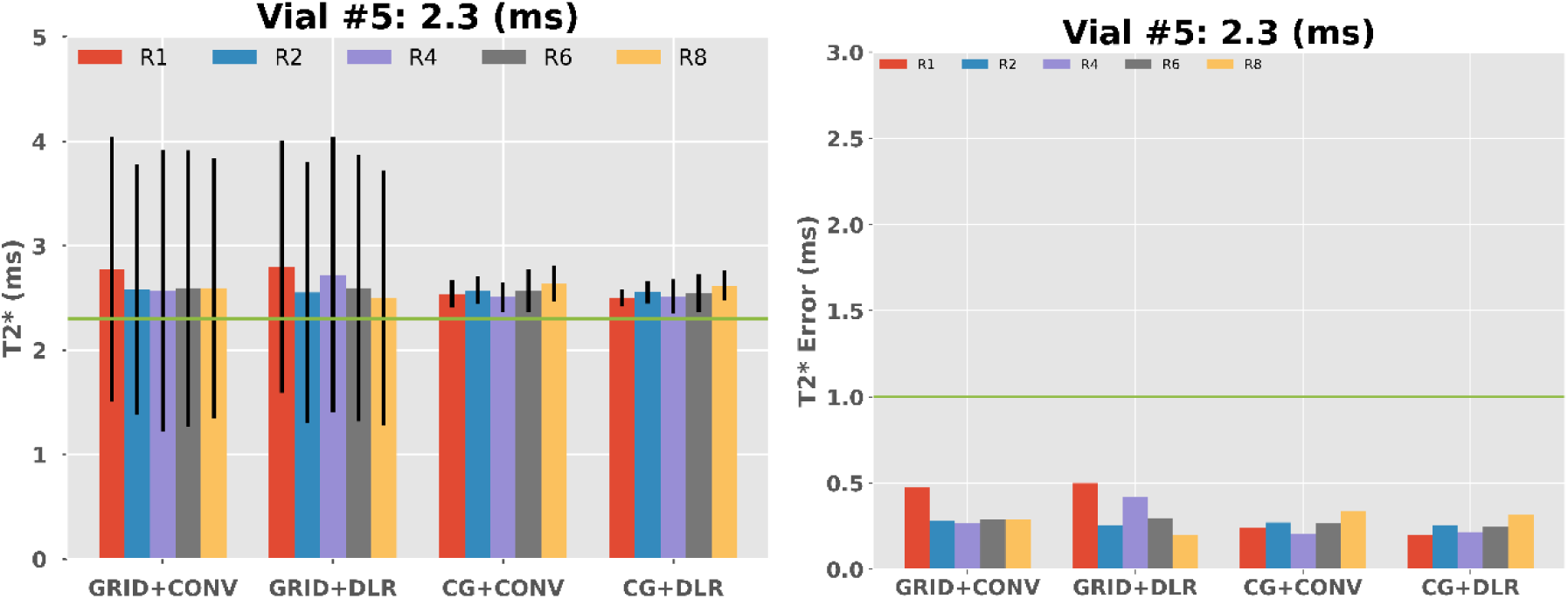
T2* (left) and T2* error (right) measured from **vial #5** with different accelerations and reconstruction methods. Error bars represent the spatial standard deviation of T2* values within an ROI.

**Supplemental Fig. 8.**
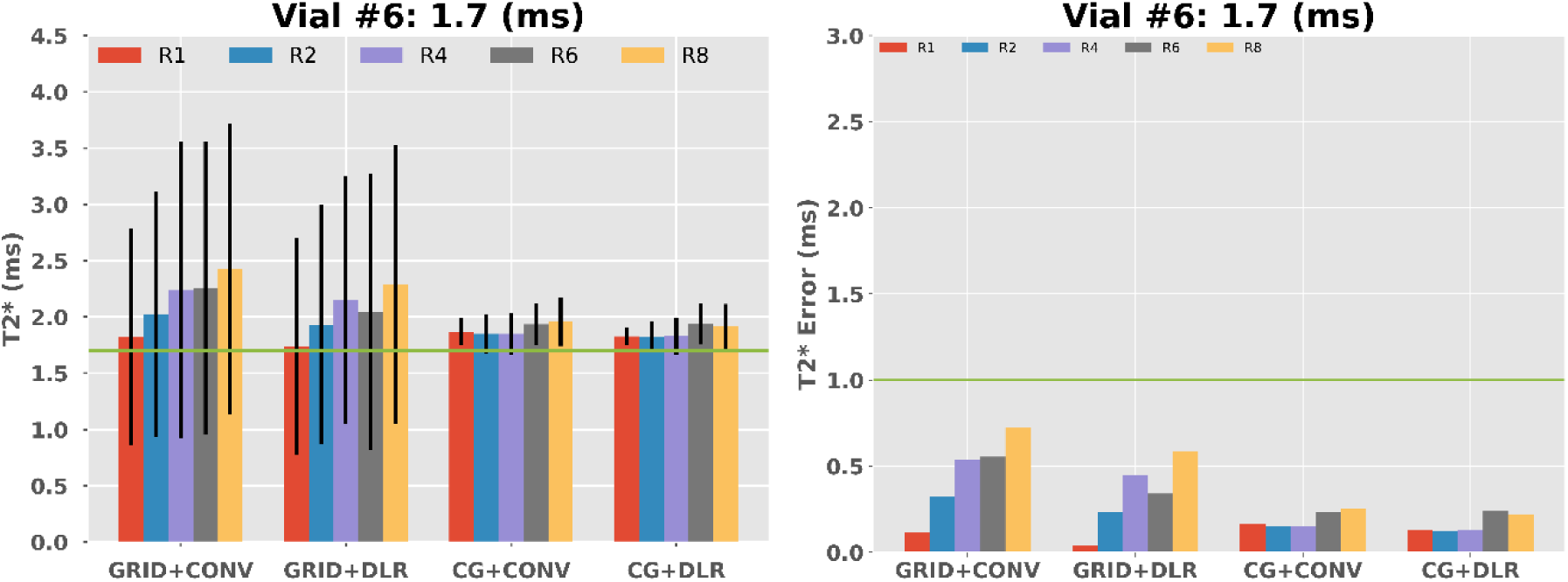
T2* (left) and T2* error (right) measured from **vial #6** with different accelerations and reconstruction methods. Error bars represent the spatial standard deviation of T2* values within an ROI.

**Supplemental Fig. 9.**
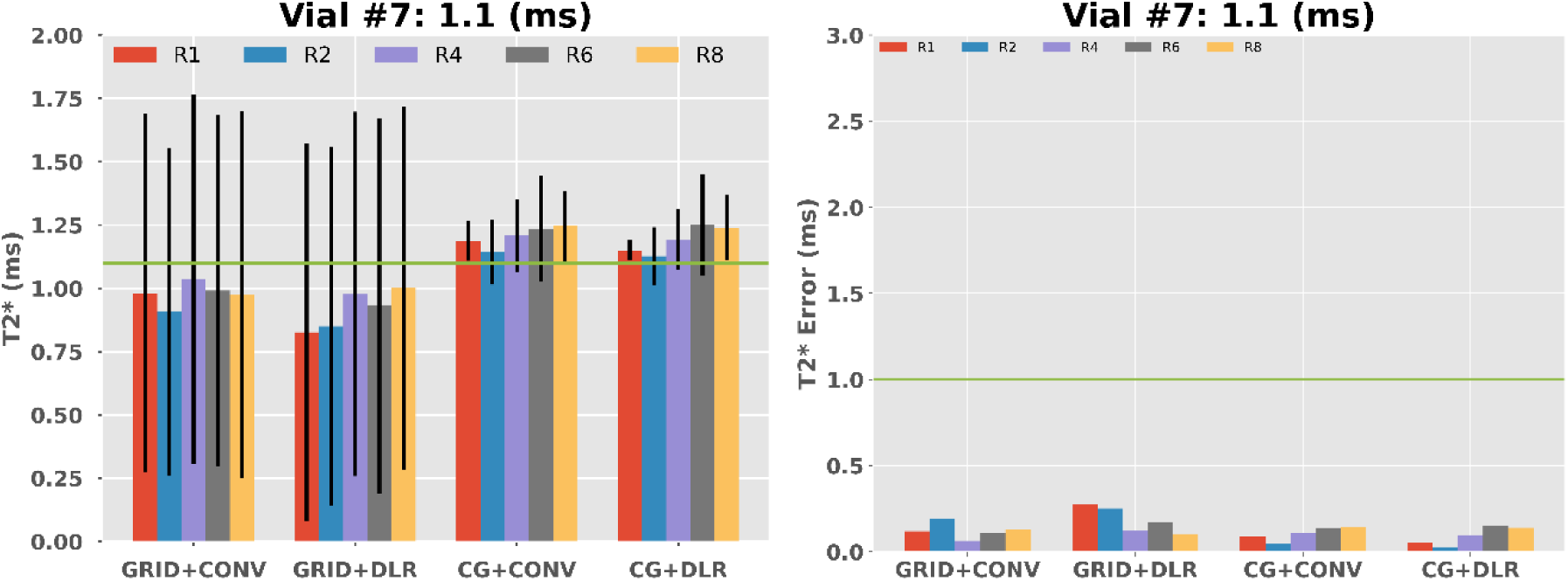
T2* (left) and T2* error (right) measured from **vial #7** with different accelerations and reconstruction methods. Error bars represent the spatial standard deviation of T2* values within an ROI.

**Supplemental Fig. 10.**
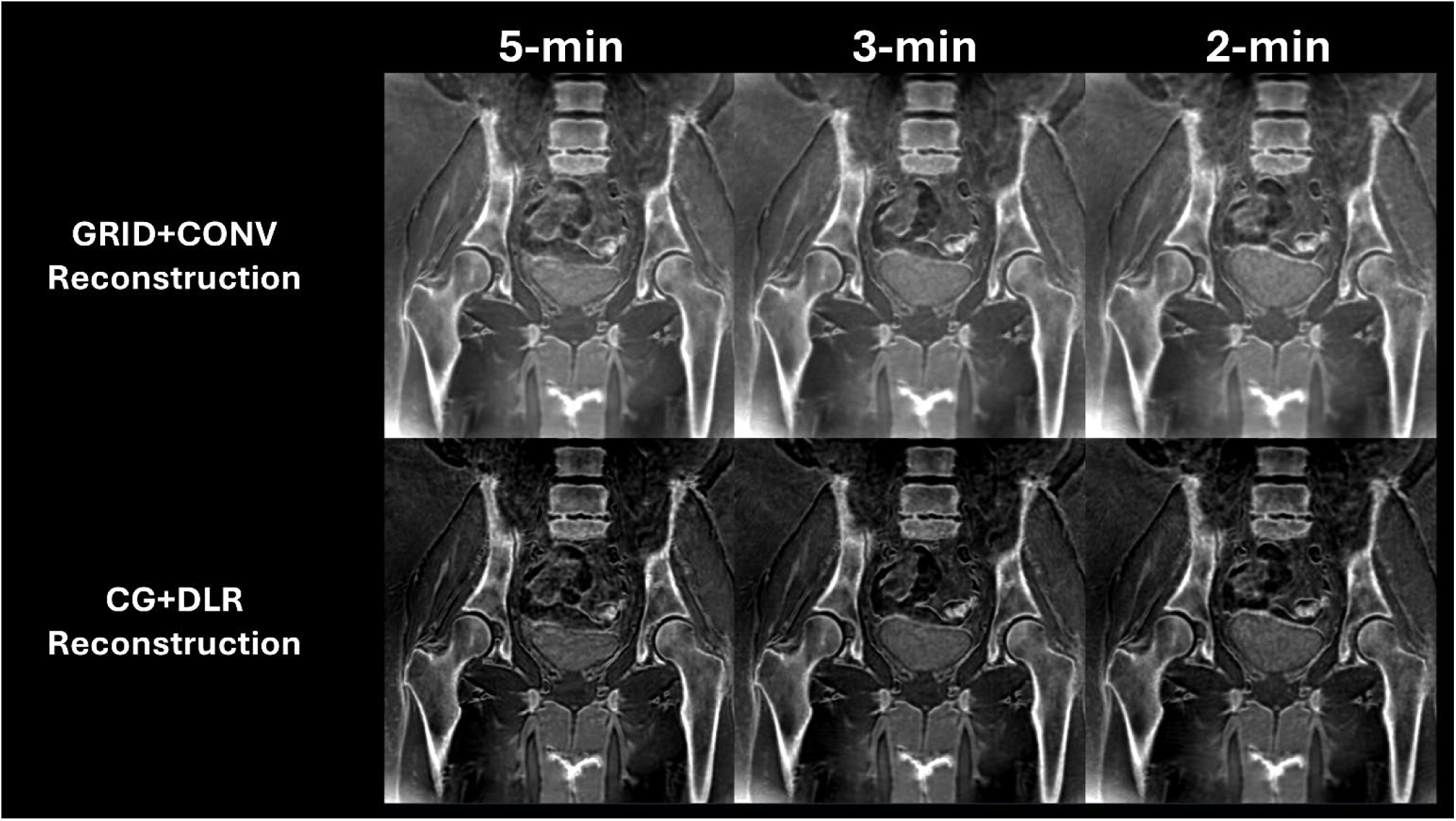
Bone-weighted images of a bilateral hip acquired prospectively and reconstructed with GRID+CONV (top row) and CG+DLR (bottom row). Qualitatively, CG+DLR provides better sharpness compared to GRID+CONV. All CT-like images are displayed on the same gray scale.

**Supplemental Fig. 11.**
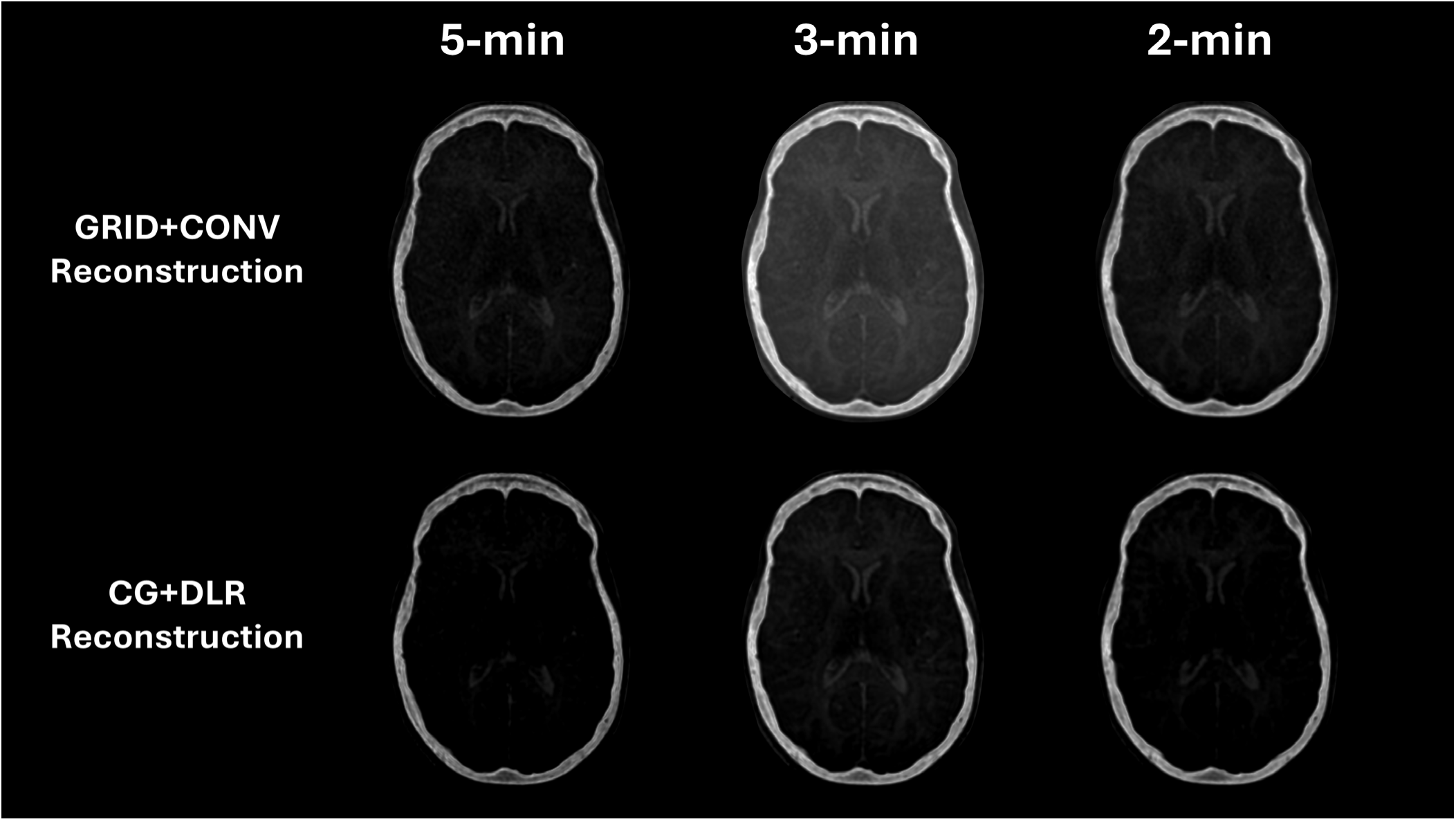
Bone-weighted images of a skull acquired prospectively and reconstructed with GRID+CONV (top row) and CG+DLR (bottom row). Qualitatively, CG+DLR provides better sharpness compared to GRID+CONV. All CT-like images are displayed on the same gray scale.

## Notes

### Summary of Updates

This version of the manuscript has been revised to update the title.

